# Stimulation better targets fast ripple generating networks in super-responders to the responsive neurostimulator system (RNS)

**DOI:** 10.1101/2022.11.30.22282937

**Authors:** Shennan Aibel Weiss, Daniel Rubinstein, John Stern, Dawn Eliashiv, Itzhak Fried, Chengyuan Wu, Ashwini Sharan, Jerome Engel, Richard Staba, Michael R. Sperling

## Abstract

**Objective:** How responsive neurostimulation (RNS) decreases seizure frequency is unclear. Stimulation may alter epileptic networks during inter-ictal epochs. Since fast ripples (FR) may be a substrate of the epileptic network, we examined whether stimulation of FR generating networks differed in RNS super- and intermediate-responders.

**Methods:** In 10 patients, we detected FR during sleep from stereo EEG (SEEG) contacts during the pre-surgical evaluation in patients with subsequent RNS placement. The normalized coordinates of the SEEG contacts were compared with that of the eight RNS contacts, and RNS stimulated SEEG contacts were defined as within 1.5 cm^3^ of the RNS contacts. We compared the post-RNS placement seizure outcome to 1) the ratio of stimulated SEEG contacts in the seizure-onset zone (SOZ SR); 2) the ratio of FR events on stimulated contacts (FR SR); and 3) the global efficiency of the FR temporal correlational network on stimulated contacts (FR SGe).

**Results:** We found that the SOZ SR (p=0.18) and FR SR (p=0.06) did not differ in the RNS super- and intermediate-responders, but the FR SGe did (p=0.02). In super-responders highly active desynchronous sites of the FR network were stimulated.

**Significance:** RNS that better targets FR networks, as compared to the SOZ, may reduce epileptogenicity more.

**Short summary:** Responsive neurostimulation (RNS) may reduce seizures by altering the epileptic network. Fast ripples (FR) may be a substrate of the epileptic network. We investigated, in 10 patients, if RNS stimulation of FR generating brain differed in RNS super-responders and intermediate-responders. The RNS stimulated brain sites were defined as contacts in the pre-surgical stereo EEG implant within 1.5 cm^3^ of the subsequently placed RNS stimulation contacts. FR events were more highly active and temporally desynchronous (p<0.05) on the stimulated contacts in the RNS super-responders. We show RNS that better targets FR networks, as compared to the seizure onset zone, may reduce epileptogenicity more.

## Introduction

The responsive neurostimulator system (RNS) is increasingly utilized by clinicians to reduce seizure frequency in patients with seizure onset zone(s) located bilaterally or in eloquent cortex^1–3^. The RNS is a closed-loop neurostimulator that responds to specific intracranial EEG (iEEG) activity. An implanted intracranial programmable neurostimulator is connected to two four contact depth electrode and/or subdural electrode leads. These leads are placed in the region of a presumed seizure onset zone(s). The neurostimulator continually measures the iEEG activity via the recording contacts and delivers electrical stimulation in response to detection of activity that differ from the usual background. The physician can easily calibrate the threshold for the detection of deviation of EEG signals and the stimulation parameters.

Placement of the RNS device was initially thought to reduce seizure frequency by stimulating during ictal epochs and aborting the seizure^4^. However, the RNS device stimulates the brain over 1000 times a day and almost entirely during the inter-ictal epoch^5^. Seizure frequency decreases gradually over years following RNS^3^. Also, RNS efficacy is better predicted by the features of stimulation during inter-ictal rather than ictal epochs^6^. Furthermore, closed- and open-loop stimulation have been shown to be similarly effective^7^. One study found that a reduction in seizure frequency, following RNS placement, is correlated with measures of coherence in the low frequency iEEG, measured over 1-3 years post implant^8^. Thus, the efficacy of RNS may be more strongly related to induced alterations in the epileptic network^8^.

We hypothesize that the RNS mechanism of action is to modulate fast ripple (FR) networks and reduce seizures because of the intrinsic role of FR in epileptogenicity and ictogenesis^9–13^. Alternatively, FR may be just a biomarker of the epileptogenic zone (EZ) that is well suited for targeting by RNS^11^. We have previously found that patients with seizure improvement after RNS placement have a less widespread FR network with less highly active desynchronized FR sites as compared to: 1) patients not offered an RNS because of widespread seizure onset zones; and 2) non-responders to resection. These results implicate modulation of FR networks in seizure reduction following RNS placement^11^.

## Methods

The institutional review board (IRB) at Thomas Jefferson University (TJU) and University of California, Los Angeles (UCLA) approved the acquisition and secondary analysis of the data used in this study. Data were collected for research purposes without impacting diagnostic studies or clinical decision-making.

This retrospective neurophysiological study used clinical iEEG recordings from 6 patients who underwent intracranial monitoring with stereo EEG (SEEG) electrodes between at UCLA and 4 patients at TJU between 2015 and 2018 for the purpose of localization of the seizure onset zone. Patients had pre-implant T1-weighted magnetic resonance imaging (MRI) for MRI-guided stereotactic electrode implantation, a post-implant CT scan to localize the depth electrodes, and a post-RNS placement CT scan to localize RNS stimulation contacts. Clinical iEEG (2000 samples per second; 0.1–600 Hz) was recorded from the SEEG contacts during sleep (Supplemental Methods). The recordings from the RNS leads were not analyzed in this study. The attending neurologist determined the seizure onset zone (SOZ) from review of video-EEG of the patient’s habitual seizures. For each patient electrodes contacts labeled as the SOZ were aggregated across all seizures and did not include areas of early propagation. Post-RNS implant outcome using the classification scheme of Khambhati in which super-responders are defined as greater than 90% reduction in seizures, and intermediate responders as a 50-90% reduction in seizures^8^. Outcome was determined using the patient reported clinical seizures at the time of most recent clinic visits compared to baseline reported clinical seizure frequency prior to RNS implantation.

Individual, electrode contacts were labeled in the post-implant and post-RNS CT and coregistered with the pre-implant MRI and then projected in to normalized MNI coordinates (Supplemental Methods). SEEG contacts stimulated by the RNS were determined as SEEG contacts within a radius of 1.5 cm from any one of the eight RNS stimulation contacts (Supplemental Methods).

High-frequency oscillations (HFOs) and sharp-spikes were detected and characterized using previously published methods implemented in Matlab (Mathworks, Natick, MA, USA)(Supplemental Methods). We selected only fast ripple on oscillations with a spectral content > 350 Hz and all fast ripple on spikes for our analysis because they have a higher propensity to be generated in epileptogenic regions^11,13^. The SOZ stimulation ratio (SOZ SR) was defined as the total number of RNS stimulated SOZ SEEG contacts in the numerator and the total number of SOZ SEEG contacts in the denominator. The FR stimulation ratio (FR SR) was defined as the total number of FR events on RNS stimulated contacts in the numerator and the total number of FR events recorded by all the SEEG contacts in the denominator. The FR stimulated global efficiency (FR SGe) was derived by calculating an adjacency matrix of the mutual information (MI) between FR spike trains, defined by the FR onset time, between stimulated and first-degree neighboring contact pairs and then using Brain Connectivity Toolbox function charpath.m (Supplemental methods).

The Mann-Whitney U test was used to compare the SOZ SR, FR SR, and RR SGe between the RNS super-responders group and the intermediate-responders group. We did not correct for multiple comparisons because we used only three planned comparisons and had a small sample size. Brainnet viewer was used for visualization (Supplementary Methods).

## Results

Three of the ten patients who were treated with RNS placement were classified as super responders (S), and the other seven as intermediate responders (I)^8^. No patients were classified as poor responders. The RNS was placed at least four years prior to outcome determination (Table 1). In all the patients all eight contacts were used for stimulation. We found no obvious relationship between the lead type or the time from RNS implantation to last follow up with RNS outcome in our small cohort (Table 1).

**Table 1:** Patient characteristics. Note RNS lead type is not bilateral unless specified. Abbreviations: L: left, R:right, LOC:loss of consciousness, AVM:arteriovenous malformation, MT:mesial-temporal

| Patient ID | Risk Factor | PET | MRI | iEEG SOZ | RNS lead type (Depth, subdural) | Years since RNS implant | Post-RNS seizure outcome |
| --- | --- | --- | --- | --- | --- | --- | --- |
| 3915 | minor head injury no LOC | Normal | R temporal | L middle temporal gyrus, MT | Depth Strip | 5 years | Super responder |
| 3394 | None | Normal | R temporal | cingulate gyrus, middle temporal gyrus, bilateral MT | Bilateral Depths | 4.5 years | Super responder |
| 4163 | Sub-arachnoid hemorrhage | Encephalo-malacia | L temporal | L pre-central gyrus | Depth Strip | 5.5 years | Intermediate responder |
| 4175 | minor head injury no LOC | Normal | N/A | R cingulate gyrus, SMA, post-central gyrus, precuneus, superior parietal lobule | Strip Strip | 5.5 years | Intermediate responder |
| 458 | None | Normal | R temporal | Bilateral MT, uncus. | Bilateral Depths | 7 years | Intermediate responder |
| 463 | AVM | R occipital AVM | R occipital | Bilateral MT, uncus. | Bilateral Depths | 6 years | Intermediate responder |
| 468 | None | L MT FLAIR | R and L temporal | bilateral MT, inferior temporal gyrus, middle temporal gyrus, fusiform gyrus | Bilateral Depths | 5 years | Super responder |
| 470 | None | L temporal lobe hypo-metabolism | L hippocampal hyper-intensity & atrophy | L MT | Depth Depth | 5 years | Intermediate responder |
| 478 | None | Peri-ventricular nodular heterotopia, hypothalamic hamartoma | Normal | bilateral MT, fusiform gyrus, superior temporal gyrus, middle temporal gyrus, inferior temporal gyrus | Depth Depth | 4.5 years | Intermediate responder |
| 481 | TBI w/o LOC | L MTS | R AND L temporal | L MT, middle temporal gyrus, inferior temporal gyrus, fusiform gyrus, uncus, inferior frontal gyrus, middle frontal gyrus | Depth Depth | 4.5 years | Intermediate responder |

We determined the pre-RNS placement SEEG contacts that could be considered as stimulated by the RNS system after the device had been placed. In our cohort, the RNS stimulation parameters had changed many times since implantation. Since stimulation parameters have been found not to strongly predictive of outcome^14^, we did not incorporate the individual most recent stimulation parameters into our calculations for identifying the stimulated contacts. Instead, we defined the pre-implant SEEG electrode stimulated contacts as within a radius of <1.5 cm (Supplemental Methods) of the eight RNS contacts (i.e., two leads of either a four-contact depth or subdural strip). Our justification for using this method, and a threshold distance of <1.5 cm, was that RNS stimulation intensities typically ranged from ∽1-3 mAmps at UCLA and TJU, and if the stimulation was monopolar, the stimulation would produce an electric field of ∽2-4 mV/mm at 1.5 cm away from the stimulation source^15^. Electric field strengths of at least 2-4 mV/mm can induce spike field coherence^16,17^, and smaller fields at distances greater than 1.5 cm may only influence spike timing^18^. Notably, a closer proximity of stimulation contacts to epileptogenic regions was required for typical RNS stimulation parameters to abort seizures^4^, but we were interested in inter-ictal effects. Our model ignores the exact placement of the contacts in gray or white matter, the stimulation train frequency and duration, and the number of stimuli delivered each day and therefore introduces considerable variability, and many confounds. However, we were most interested in comparing the relative proximity of RNS stimulation contacts to FR networks as compared to the SOZ.

After designating the SEEG contacts stimulated by RNS, we compared whether the proportion of stimulated SOZ contacts (SOZ stimulation ratio [SR]), and proportion of FR generated on the stimulated contacts (FR SR) differed for the RNS super-responder patients. Next, by comparing the FR event timing across the stimulated contacts and their first-degree neighbors, we asked if the FR temporal synchrony that defines the FR network would differ in the super-responders. In a prior study, we had found that temporally desynchronous nodes of the FR network also generate FR at higher rates and, if left unresected, correlate with a non-seizure free outcome^13^. Therefore, we hypothesized that in the super responders the stimulated FR networks would be more desynchronous. To measure the synchrony of the stimulated FR network we used the FR stimulated global efficiency (FR SGe).

We found that the 3 super-responders exhibited no difference in the SOZ SR compared to the intermediate-responders (p=0.18, Figure 1A1,B). The FR SR measured using FR detected during sleep in the SEEG implant trended higher in the super-responders (p=0.06, Figure 1A2,1C) indicating a less widespread FR network that is better targeted by RNS stimulation. In the super-responders the FR SGe was significantly lower (p=0.02, Figure 1A3,1D) indicating that in the three super responders the RNS leads were targeting more desynchronized sites in the FR network^13^, as compared to the intermediate-responders. In summary, results show that the three super-responders exhibited significantly more proximity of the RNS stimulation contacts to desynchronous, and highly active^13^, sites in the FR network in comparison to the full extent of the SOZ.

**Figure 1:**
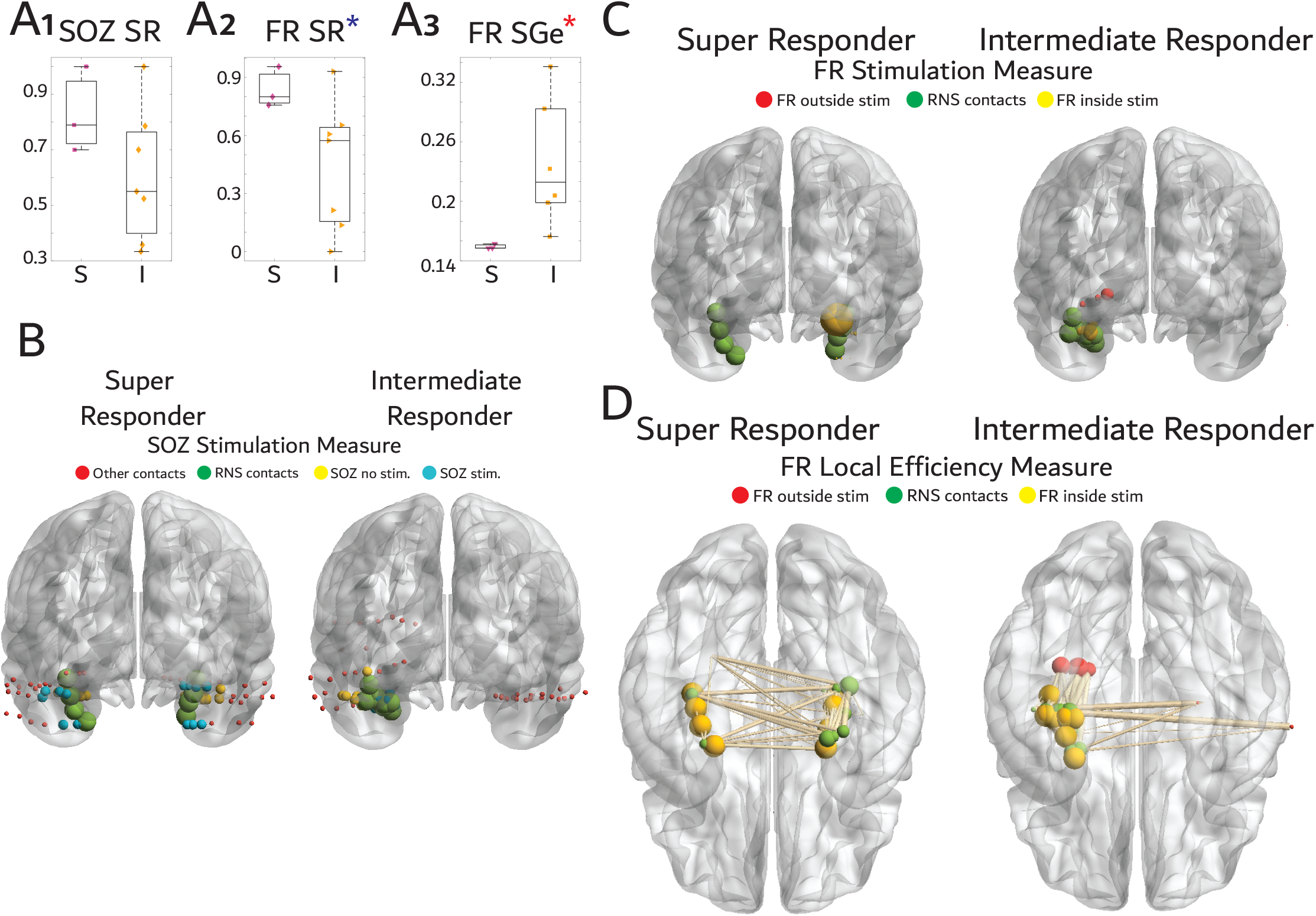
Stimulation targets fast ripple (FR) generating networks, that are highly active and desynchronous, in super responders to the responsive neurostimulator system (RNS). (A) Box plots of the seizure onset stimulation ratio (A1, SOZ SR), FR stimulation ratio (A2, FR SR), and FR stimulated global efficiency (A3, FR SGe) stratified by RNS seizure outcome of super-responder (S) and intermediate-responder (I). Black asterisk indicates a trend (p=0.06), red asterisk indicated statistical significance (p<0.05). (B) Illustrative brain graph of the SEEG and RNS implants in a super-responder (left) and intermediate-responder (right). The RNS contacts are colored green, the stimulated SOZ contacts are cyan, the non-stimulated SOZ contacts are colored yellow, other contacts are colored red. (C) Illustrative brain graph as in B but only SEEG contacts generating FR are included. FR generating contacts (*i.e*., nodes) that are stimulated are colored yellow, unstimulated FR generating contacts are colored red. The size of the red and yellow nodes is proportional to the FR rate of that node. (D) Illustration of the FR mutual information network used to calculate the FR SGe. The nodes colored yellow are stimulated, and the nodes colored red are unstimulated. The size of the red and yellow node correspond to the nodes local efficiency. The edges are weighted by the FR mutual information value. Note that in C and in D the intermediate-responder has more unstimulated FR nodes, and stimulated nodes with a high local efficiency and higher mutual information edges as compared to the super-responder, respectively.

## Discussion

RNS may derive its efficacy in reducing seizure burden by inducing neuroplastic changes in epileptic networks over months and years^3,6–8^. Epileptic networks are theoretical concepts and can be derived in a multitude of ways such as coherence changes in lower-frequency EEG rhythms measured by the patient’s RNS system^8^. Fast ripples are known to be important in epileptogenesis, the generation of epileptiform discharges, and seizure genesis and could potentially be one of the more important substrates of the epileptic network^11–13^. Our results show that RNS super-responders had the most active and desynchronous sites of their FR network targeted by the RNS stimulation electrodes. However, since this study did not establish causality, FR networks may merely be the biomarkers of the EZ^11^, which when targeted by RNS improves seizure frequency reduction irrespective of modulation of FR generation.

FR are thought to be generated by microcircuits known as pathologically interconnected neurons (PIN) clusters^9^. It is conceptualized that in epileptic networks these PIN clusters act as internal kindling generators that recruit additional territories over months and years and promote seizures on a shorter time scale^9^. The individual FR events are known to propagate from one region to another^10,12^ and have been associated with the generation of epileptiform spikes^12^. Potentially, stimulation by the RNS system could reduce the probability of activation of these PIN clusters and reduce seizure probability^9^. Further work investigating the effects of closed- and open-loop neurostimulation in the experimental epilepsies is required to further examine this hypothesis.

Our study had several important limitations. As noted in the results, our methodology for deriving the stimulated electrodes was greatly simplified. Electrical fields from a monopolar source decrease proportional to the inverse square of the distance, and furthermore whether these fields induce spike-field coherence or not is not an absolute. Therefore, at the very least, our results imply only that FR generating sites and networks are more proximal to RNS stimulation contacts in super-responders, as compared with the SOZ contacts. Additionally, our study cohort was very small, and these results need to be repeated in a larger cohort. Based on our sample size, a much larger cohort is required to prove statistical superiority of the FR SR or FR SGe as compared to the SOZ SR. Future works should also utilize statistical models that account for other factors, such as the location of the SOZ and the baseline seizure frequency, that can also potentially influence RNS seizure outcome. Finally, all the epileptogenic regions may not have been spatially sampled by the SEEG implant which would impact interpretation of all three SEEG derived measures.

In conclusion, our small study shows that, in RNS super-responders as compared to RNS intermediate-responders, the RNS stimulation electrodes are more proximal to highly active desynchronous FR network sites as compared to the SOZ. More work is needed to determine if RNS should target FR networks as opposed to SOZ sites, and if RNS modulates FR networks or rather FR serve solely as a better biomarker of the EZ.

## Data Availability

All data produced in the present study are available upon reasonable request to the authors

## Acknowledgements

The authors thank Mr. Kirk Shattuck, Dr. Iren Orosz, and Mr. Dale Wyeth and Mr. Ted Wyeth for their technical contribution in collecting EEG recordings, and Dr. Iren Orosz and Dr. Richard Gorniak for assistance with the neuroimaging data.

