## supplementary methods for "Stimulation better targets fast ripple generating networks in super-responders to the responsive neurostimulator system (RNS)"

Supplemental Methods

iEEG recordings: For each patient, clinical iEEG (0.1–600 Hz; 2000 samples per second) was recorded from 8 to 16 depth electrodes, each with 7–15 contacts, using a Nihon-Kohden 256-channel JE-120 long-term monitoring system (Nihon-Kohden America, Foothill Ranch, CA, USA). A larger number of electrodes with more contacts were implanted at TJU. The reference signal used for the recordings performed at UCLA was a scalp electrode position at Fz in the International 10–20 System. The reference signal used for the TJU recordings was an electrode in the white matter that was: 1) estimated far from the cortical layers based on the electrode design and trajectory; 2) far from the putative SOZ based on scalp recordings; and 3) exhibited low voltages relative to the other contacts on the electrode.

Neuroimaging and derivation of stimulated contacts: A custom pipeline (<https://github.com/pennmem/neurorad_pipeline>) was used to derive the normalized MNI coordinates of the SEEG contacts and the RNS contacts^1–3^. Voxtool (https://github.com/pennmem/voxTool) was used on the post-SEEG implant CT and post-RNS placement CT to label the SEEG contacts and RNS stimulation contact, respectively. FreeSurfer (http://surfer.nmr.mgh.harvard.edu/) was used on the MRI to construct individual subject brain surfaces and cortical parcellations according to the Desikan–Killiany atlas^4^. The Advanced Neuroimaging Tools^5^ was used to separately coregister: 1) the post-SEEG implant CT with the MRI; and 2) the post-RNS placement CT with the MRI and the position of each electrode contact was localized to the Desikan-Killiany atlas. The pipeline was then used to transform the position of each electrode contact from individual subject space to an averaged FreeSurfer space with normalized Montreal Neurological Institute (MNI) coordinates (defined by the fsaverage brain).

To determine the stimulated SEEG contacts the Euclidian distance calculated as $d=\sqrt{{(X_{RNS}-X_{SEEG})}^{2}+{(Y_{RNS}-Y_{SEEG})}^{2}+{(Z_{RNS}-Z_{SEEG})}^{2}}$ was derived between all the RNS and SEEG electrodes. SEEG with a single Euclidian distance value for a RNS and SEEG pair less than or equal to 15 mm in Normalized MNI coordinates were designated stimulated electrodes. The threshold of 15 mm was selected according to the equation $\frac{dV}{dr}=\frac{I}{4\pi{\sigma r}^{2}}$ which derives the electric field strength produced by a monopolar current source, where V is voltage, I is current, r is the distance from the monopolar source, and σ is the conductivity of gray matter^6,7^.

HFO detection

In brief, the HFO detector reduced muscle and electrode artifacts in the iEEG recordings using an independent component analysis (ICA)-based algorithm^8^. After applying this ICA-based method, ripples and fast ripples were detected in the referential and bipolar montage iEEG recordings per contact by utilizing a Hilbert detector, in which a 1,000th order symmetric finite impulse response (FIR) band-pass filter in the 200-600 Hz band for fast ripples was applied, and (ii) a Hilbert transform was applied to calculate the instantaneous amplitude of this time series according to the analytic signal z(t) in Eqn. 1.

z(t)=a(t) e^(iθ(t))

where a(t) is the instantaneous amplitude and θ(t) is the instantaneous phase of z(t). Following the Hilbert transform, the instantaneous HFO amplitude function [a(t)] was smoothed using moving window averaging, the smoothed instantaneous HFO amplitude function was normalized using the mean and standard deviation of the time series, and a statistical threshold defined by the skewness of the normalized band pass filtered time series was used to detect the onset and offset of discrete/potential events^9–13^.

Selection of a referential or bipolar montage for individual channels or channel pairs, within patients, was based on either visual inspection or automatically selected by the detector. In the former case, patients recorded with Nihon Kohden only have grounding to the first 64 channels and thobe beginning 65 and higher are not, and thus a bipolar montage best reduces noise.In the latter case, machine learning based on several statistical features of the signal were used to automatically transition referential channels to bipolar pairs.

HFO-like events can arise due to Gibb’s phenomenon, i.e., high-pass filtering sharp transients, including epileptiform spikes^14^. To distinguish authentic HFOs from authentic HFOs on EEG spikes or spurious HFO due to filter ringing, we used an algorithm that performed topographic analysis of time-frequency plots for each HFO and defined open- and closed-loop contour groups^9,15^. The algorithm also measured the power, spectral content, duration, onset time, and offset time of each HFO and categorized the HFO as an HFO on oscillation, HFO on spike, or sharp-spike (i.e. false HFO). HFOs that did not coincide with spikes were defined as closed-loop contour groups with an onset that did not overlap with the onset of open-loop contour groups. For HFOs on spikes the two groups temporally overlapped3,45. Following automatic detection of HFO and sharp-spikes, false detections of clear muscle and mechanical artifact were deleted by visual review in Micromed Brainquick (Venice, Italy). The complete open-source HFO detector can be downloaded at <https://github.com/shenweiss>.

Derivation of fast ripple stimulated global efficiency

The fast ripple stimulated global efficiency (FR SGe) was calculated using the Brain Connectivity Toolbox (https://sites.google.com/site/bctnet/)^16^ function charpath.m. The adjacency matrix for the mutual information (MI) networks were calculated using FR event ‘spike trains’ defined by the onset times of each event and then calculating MI between electrode contacts (*i.e.,* nodes) with the adaptive partition using inter-spike intervals MI estimator^17^. To calculate the distance the inverse of the adjacency matrix was derived. To calculate the FR SGe, edges between non-stimulated nodes (i.e. non-stimulated node:non-stimulated node) were assigned an infinite distance.

Data visualization

Example weighted node distributions and examples of FR MI networks were rendered with Brainnet viewer^18^.

7. Plonsey R, Barr RC. Bioelectricity A Quantitative Approach. Kluwer Academic/Plenum Publishers; 2000.
